# Productivity losses and treatment cost of Benign Prostatic Hyperplasia in Ghana

**DOI:** 10.1101/2025.03.18.25324214

**Authors:** Daniel Senanu Dadee-Seshie, Sedem Godson Agbeteti, Banabas Kpankyaano, Aishah Fadila Adamu, Selase Kofi Adanu, Yakubu Alhassan, Kekeli Kodjo Adanu

## Abstract

Benign Prostatic Hyperplasia (BPH) is a common condition among aging men, characterized by lower urinary tract symptoms that significantly impact quality of life and economic productivity of affected individuals. The financial burden of BPH extends beyond direct medical expenses to include direct non-medical costs and productivity losses. In Ghana, limited data exist on the cost implications of BPH, leaving a critical gap in healthcare planning and resource allocation.

We aimed to determine the cost and productivity losses associated with diagnosing and managing BPH at the Ho Teaching Hospital.

A cross-sectional cost-of-illness study was conducted at the Urology Unit of Ho Teaching Hospital from July to September 2024. Data were collected from 105 patients diagnosed with BPH using structured questionnaires. Direct medical costs, including consultation, diagnostics, medication, and surgical interventions, were calculated using a bottom-up approach. Direct non-medical costs covered transportation, food, lodging, and caregiver expenses, while productivity losses were estimated based on absenteeism and reduced working hours, using the human capital approach.

The mean monthly direct medical cost per patient was GHS 5,464.30 ($370.46), with surgeries accounting for 30% of total expenses. The total direct non-medical costs, dominated by transportation (66.3%), amounted to GHS 9,842.00 ($667.25). Productivity losses due to absenteeism and caregiving responsibilities totaled GHS 4,746.28 ($321.78), with 30% of employed patients missing work. Notably, direct medical costs contributed the highest economic burden (86.5%), surpassing direct non-medical costs (9.1%) and productivity losses (4.4%).

BPH imposes a significant financial burden on patients and households in Ghana, driven by high out-of-pocket medical costs, non-medical expenses, and lost productivity. The findings underscore the need for cost-effective treatment strategies, improved health insurance coverage, and targeted interventions to alleviate financial hardships associated with BPH management.

## Introduction

Benign prostatic hyperplasia (BPH) is a common condition affecting elderly and middle-aged men, characterized by the enlargement of the prostate gland, which can lead to urinary symptoms [1]. The severity and combination of the symptoms can vary across individuals. Common symptoms include urinary frequency, nocturia, hesitancy, and poor urinary stream among others. BPH affects millions of men worldwide and presents a significant public health concern [2]. The global prevalence of BPH varies significantly across different regions and populations. Studies indicate that approximately 26.2% of men will experience BPH in their lifetime, with prevalence rates increasing with age [3]; and in Africa, prevalence rates ranging from 6.1% to 33.4% have been reported [4]. A study conducted in Ghana, observed that 48.5% of participants aged 40 years and above had BPH indicating a significant presence of the disease in the Ghanaian population [5]. Coupled with the high disease burden is the productivity losses and the cost implications of the disease.

One of the areas impacted is work productivity [6], which is lost because of absenteeism and presenteeism among employees with the illness. The associated lower urinary tract symptoms affect productivity in terms of the reduced hours spent at work compared to when they had no symptoms. Meanwhile, the cost of treating BPH has been on the ascendency. Studies conducted by Scott and Scott [7] in New Zealand marked an early effort to estimate the costs of treating BPH in both public and private sectors. By considering hospital data, the study reported the annual total direct medical costs for treated BPH in 1991 at $NZ16 million. A more recent study estimated the annual direct medical cost of BPH to the UK economy at GBP 180.8 million [8]. Due to socio-economic factors, change in demography and innovations in surgery, the cost of treatment is likely to soar. An aging population and rise in life expectancy are also expected to further worsen the cost of care.

In the US (United States), significant annual costs are reported for medications and diagnostic tests [9]. Long-term medications for BPH are more expensive than surgery [10]. Direct non-medical costs, such as informal care, transportation, and specialized products like absorbent pads, significantly contribute to the overall economic burden of BPH [11]. In Hungary, these costs account for 31% of the total treatment cost (€876) [12]. Rural patients face high transportation costs [13], and caregiver expenses add further financial strain [14]. Dietary changes also impose costs, though specific figures are not well-documented [15].

Productivity loss due to BPH significantly impacts the economy, costing the US an estimated 500 million out of the total annual cost of BPH ($3.9 billion)[8]. BPH interferes with daily activities [8,16] and patients experience low physical activity, absenteeism, and presenteeism, affecting work efficiency [12,17]. Despite its prevalence, comprehensive research on BPH-related productivity loss is sparse [18].

Literature on the treatment cost of BPH in Ghana is severely limited and to the best of our knowledge, there is no comprehensive evidence on the subject. Yeboah [19] reported that the cost of BPH medications ranged from USD 300 – 500 while the cost of simple prostatectomy/Transurethral resection of the prostate (TURP) was estimated at USD 1100. These estimates were however anecdotal at best with no empirical data to buttress it. Due to this lacuna, relevant data to inform healthcare planning and resource allocation is lacking. In addition, expenditure on BPH is not only of public health concern because the household economy is equally implicated. Families make substantial out-of-pocket payments to defray the cost of care thereby forcing them to sacrifice other basic needs, sell assets and eventually become impoverished [20]. Thus, BPH treatment may have consequences on patients’ quality of life and well-being; and to avert catastrophic health expenditure, interventions must be undergirded by empirical data.

To this end, this study determined the direct medical cost, indirect medical costs and productivity losses of Benign Prostatic Hyperplasia at a tertiary health facility in Ghana.

## Materials and Methods

### Ethical statement

Ethical approval was obtained from the UHAS Research Ethics Committee with certificate number: UHAS-REC A.9 [44] 23-24. A written informed consent was obtained from all study participants.

### Study design

This study used a cost-of-illness cross-sectional approach to collect data from the patients’ perspective on the cost and productivity losses of Benign Prostatic Hyperplasia at the Ho Teaching Hospital. A total of 105 participants were recruited from 11^th^ June 2024 to 30^th^ September 2024.

### Study setting

The study was conducted at the Urology unit of the Ho Teaching Hospital. This units runs an outpatient clinic that attends to an average of thirty (30) patients per day. Men diagnosed as having BPH and willing to participate were recruited.

### Sampling

To estimate the sample size, we relied on a previous study by Rencz et al [12] which looked at the cost of treating BPH in Hungary. We relied on this study due to the paucity of comprehensive data on the subject matter from Africa. This study was designed to consecutively select 99 patients, assuming a 95% confidence level and a 5% non-response rate, a standard deviation of 1829 and an error margin of 370 around the annual cost of BPH treatment.

Patients diagnosed of BPH, undergoing treatment at the Urology unit of the Ho Teaching Hospital, who consented to participate were included in this study while patients who have been diagnosed for less than 3 months were excluded.

### Methods of data collection

This study used a quantitative data collection approach. The structured questionnaire comprised of three main sections: the sociodemographic characteristics of participants, the direct cost of treatment and productivity losses. The direct medical cost was measured using a bottom-up approach, which involved quantifying each patient’s medical expenses for BPH diagnosis and treatment in a month. This included costs for doctor visits, diagnostic exams, prescription drugs, surgeries, and other related healthcare services. The direct non-medical cost comprised of elements such as transportation costs, caregiver fees, and other related expenditures per month. Productivity loss was estimated in terms of job loss due to BPH complications, absenteeism (days of work missed due to BPH-related symptoms), and time spent by family members or caregivers accompanying and caring for BPH patients during clinic visits.

Patients were consecutively recruited at the urology clinic after assuring them of confidentiality, voluntary withdrawal and data security. Questionnaires were administered through face-to-face interviews in privacy until the required number of patients was attained. Data was cleaned, validated and entered daily. Quality control measures instituted include pre-testing of questionnaires, training of research assistants and correction of errors before data entry.

### Data Analysis Direct medical cost

The direct medical cost included costs for doctor visits, diagnostic exams (for instance PSA testing, ultrasound), prescription drugs (e.g alpha-blockers, 5 alpha-reductase inhibitors), surgeries (Transurethral resection of the prostate, open prostatectomy) and other related healthcare services. These expenses were summed up to determine the total direct medical cost and to obtain the average cost per patient, it was divided by the total number of respondents.

### Direct non-medical cost

The direct non-medical cost was determined by summing up the cost of transportation, food and drinks, caregiver fees and fees for lodging/renting. The transportation cost was calculated by summing up the cost of travel by patient and caregiver to and from the hospital. Similarly, the cost of food and drinks was determined by the summation of expenses related to food and drinks while visiting the hospital. The fees for lodge/rent and caregiver fees were also tabulated. The total direct non-medical cost was estimated by adding up the various cost components. The average cost per patient was obtained by dividing the total cost by the number of respondents.

### Productivity losses

Productivity losses associated with the management of BPH was measured in terms of lost hours and absenteeism (days of work missed due to BPH-related symptoms), and time spent by family members or caregivers accompanying and caring for BPH patients in the hospital during clinic days at the expense of working hours. The number of workdays missed or productivity reduced was quantified and translated into monetary terms using the human capital approach; which estimates the value of lost productivity in terms of wages. Using the human capital approach, lost hours were multiplied by the minimum wage in Ghana which is currently GHS 18.15/day (USD 1.16/day).

### Statistical analysis

The data generated was entered into Excel 2021 and exported to SPSS version 27 for analysis. Descriptive statistical analysis was done using frequency distributions, percentages, means ± standard deviations (SD) and presented in tables and charts.

## Results

### Socio-Demography Status

The descriptive statistics of the respondents are presented in Table 1. The average age of the participants was 71.0 years (± 8.48). The largest age group was 70-79 years (40.95%), followed by 60-69 years (29.52%), with a very small percentage (0.95%) being over 90 years old. The majority of participants were married (71.43%), while 26.67% were widowed, and 1.9% were divorced. Regarding education, 42.86% had tertiary education, and only 5.71% had no formal education. Employment status showed that 63.81% were unemployed, 17.14% were self-employed, and the rest were employed in either the private (11.43%) or public sectors (6.67%). Out of 105 participants, 40 were currently employed, while 65 were retired. Diagnosis was predominantly done by urology specialists (81%), with the remaining 19% diagnosed by primary care physicians, including house officers, medical officers, and surgical residents.

**Table 1:** Sociodemographic characteristics of study participants.

| Category | Frequency | Percentage (%) |
| --- | --- | --- |
| **Age (n=105)** |  |  |
| 45-59 | 11 | 10.48 |
| 60-69 | 31 | 29.52 |
| 70-79 | 43 | 40.95 |
| 80-89 | 19 | 18.1 |
| 90+ | 1 | 0.95 |
| Mean age ± SD | 71.0 ± 8.48 | |
| **Marital Status (n=105)** |  |  |
| Married | 75 | 71.43 |
| Widowed | 28 | 26.67 |
| Divorced | 2 | 1.9 |
| **Educational status (n=105)** |  |  |
| Tertiary | 45 | 42.86 |
| SHS | 29 | 27.62 |
| JHS/Middle School | 15 | 14.29 |
| Primary | 10 | 9.52 |
| No Education | 6 | 5.71 |
| **Current Employment Status (n=105)** |  |  |
| Unemployed | 65 | 63.81 |
| Self-Employed | 20 | 17.14 |
| Private Sector Employee | 12 | 11.43 |
| Public Sector Employee | 7 | 6.67 |
| Self employed | 1 | 0.95 |
| **Type (n=40)** |  |  |
| Business/Petty trading | 29 | 72.5 |
| Civil Service | 8 | 20 |
| Security services | 3 | 7.5 |
| **Still working or not (n=40)** |  |  |
| Yes | 40 | 100 |
| No | 0 | 0 |
| **Monthly Income (n=105)** |  |  |
| Less than \$34 | 44 | 41.9 |
| \$34 – \$135 | 41 | 39 |
| \$136 – \$338 | 11 | 10.5 |
| \$339 – \$678 | 7 | 6.7 |
| More than \$678 | 2 | 1.9 |
| **Active NHIS (n=105)** |  |  |
| Yes | 105 | 100 |
| No | 0 | 0 |
| **Physician Type (n=105)** |  |  |
| Urologist | 85 | 81 |
| Primary Care Physician | 20 | 19 |

### Direct Medical Cost Associated with the Diagnosis and Management of BPH

The mean direct medical cost was estimated at GHS 5,464.30 ($370.46), with diagnostic tests averaging GHS 278.80 ($18.90). The mean medications cost was GHS 200.74 ($13.61) and laboratory investigations (serum prostate specific antigen (PSA), full blood count, other relevant investigations specific to some patients) averaged GHS 79.14 ($5.37) per patient. Imaging studies (abdominopelvic ultrasound scan and other relevant investigations) costed GHS 52.29 ($3.55) and surgeries, GHS 4,633.33 ($314.12) per patients. No costs were reported for consultations since it is covered by the National Health Insurance Scheme. Overall, the total direct medical cost per participant averaged GHS 5,244.30 ($355.55), reflecting the diverse diagnostic approaches and medication needs among the study group. Further details are found in Table 2.

**Table 2:** Direct medical cost.

| Types of costs | Total<br>GHS(USD) | Mean Cost<br>GHS(USD) | Standard<br>deviation | Median Cost<br>GHS (USD) |
| --- | --- | --- | --- | --- |
| **Direct medical** |  |  |  |  |
| Diagnostic tests (PSA+ TRUS+ Uroflow) [n=97] | 27,044.00<br>(1,833) | 278.80<br>(18.90) | 145.04<br>(9.83) | 237.00<br>(16.07) |
| Medications | 21,078.00<br>(1,429) | 200.74<br>(13.61) | 225.30<br>(15.27) | 160.00<br>(10.85) |
| Lab Investigation | 8,310.00<br>(563.34) | 79.14<br>(5.37) | 71.03<br>(4.82) | 85.00<br>(5.76) |
| Imaging Studies | 5,490.00<br>(372.23) | 52.29<br>(3.55) | 67.13<br>(4.55) | 50.00<br>(3.39) |
| Surgeries | 27,800.00<br>(1,884.76) | 4,633.33<br>(314.12) | 1,883.26<br>(127.68) | 4,400.00<br>(12,768) |
| Other costs | 3,740.00<br>(253.56) | 220.00<br>(14.92) | 477.69<br>(32.39) | 50.00<br>(3.39) |
| Total direct medical cost | 93,462.00<br>(6,336.41) | 5,464.30<br>(370.46) | 2,869.45<br>(194.54) | 4,983.00<br>(337.88) |

\*USD 1= GHS 14.75, Ghana interbank exchange rate, July 2024

### Distribution of direct medical cost

Fig 1 provides a general overview of the various direct cost elements. Surgery accounted for 30% of the total direct cost build up while diagnostic test and medications made up 29% and 22% respectively. ‘Other’ cost component includes the cost of catheter and gels which constituted 4% of the direct medical cost.

**Fig 1:**
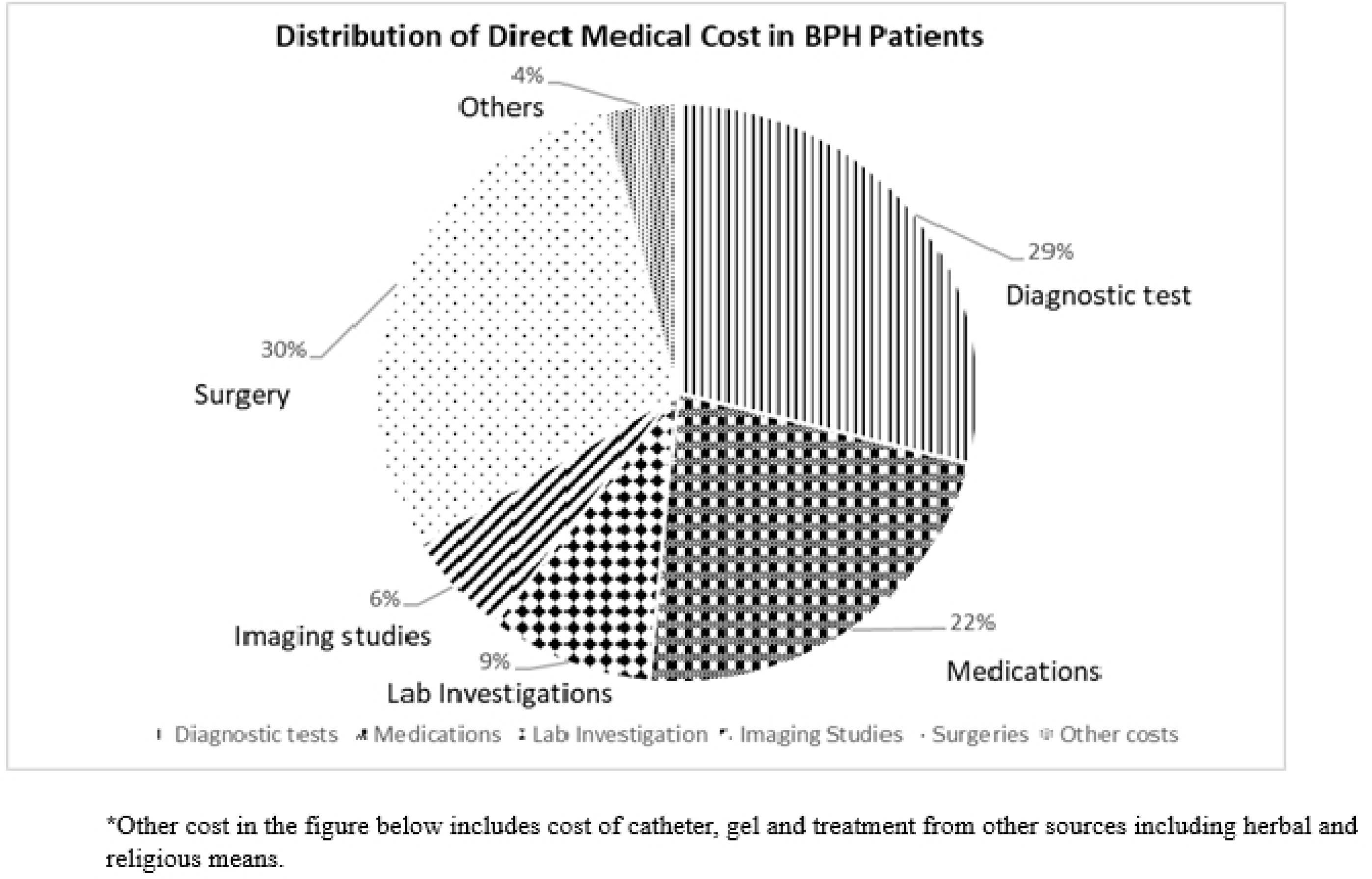
Distribution of direct medical cost.

Table 3 shows the direct non-medical costs associated with benign prostatic hyperplasia. They show the direct non-medical cost of BPH in categories: transportation, food and drinks, lodging/rent, home care services, and other non-medical costs. The total direct non-medical cost amounts to GHS 9, 842.00 ($667.25), with transportation being the highest individual cost at GHS 6,523.00 ($442.24), followed by food and drinks at GHS 3,029.00 ($205.36). Lodging/rent and home care services have significantly lower costs at GHS 200.00 ($13.56) and GHS 90.00 ($6.10), respectively.

**Table 3:** Direct non-medical cost.

| Type of cost | Total<br>GHS (USD) | Mean<br>GHS(USD) | Std. Deviation<br>GHS(USD) | Median<br>GHS (USD) |
| --- | --- | --- | --- | --- |
| **Direct non-medical cost** |  |  |  |  |
| Transportation per visit | 6,523.00<br>(442.24) | 62.12<br>(4.21) | 69.62<br>(4.72) | 40.00<br>(2.71) |
| Food and drinks (n=91) | 3,029.00<br>(205.36) | 33.29<br>(2.26) | 85.00<br>(5.76) | 20.00<br>(1.36) |
| Lodging/rent (n=1) | 200.00<br>(13.56) | 200.00<br>(13.56) | 0.00 | 200.00<br>(13.56) |
| Home care services (n=2) | 90.00<br>(6.10) | 45.00<br>(3.05) | 21.21<br>(1.44) | 45.00<br>(3.05) |
| **Total direct non-medical cost (GHS/USD)** | **9,842.00**<br>**(667.25)** | **340.41**<br>**(23.09)** | **175.83**<br>**(11.92)** | **305.00**<br>**(20.68)** |

\*USD 1= GHS 14.75, Ghana interbank exchange rate, July 2024

### Productivity Loss Associated with the Diagnosis and Management of BPH

Table 4 shows the indirect costs associated with the diagnosis and management of BPH. It details various metrics, including days absent from work, hours spent traveling and waiting before consultation, reduction in working hours, and time spent by relatives on care and transportation. The total cost of productivity loss is GHS 4,746.28 ($321.78), with a mean of GHS 195.85 ($13.28) and a standard deviation of GHS 165.06 ($11.19). The median productivity loss is GHS 145.29 ($9.85). Notable figures include a total of GHS 2,069.10 ($140.28) lost due to absenteeism from work, and significant hours spent on activities such as waiting for consultation (321.41 hours) and household care (214.94 hours). The data provide a comprehensive overview of the indirect costs incurred by BPH patients and their families.

**Table 4:** Productivity Losses Associated with BPH.

|  | **Total hours** | **Total GHS (USD)** | **Mean GHS (USD)** | **Std. Deviation** | **Median GHS (USD)** |
| --- | --- | --- | --- | --- | --- |
| Days absent from work (n=12) | 912 | 2,069.10<br>(140.28) | 137.94<br>(9.35) | 132.63<br>(8.99) | 108.90<br>(7.38) |
| Hours Spent Traveling | 175.27 | 397.86<br>(26.97) | 3.79<br>(0.26) | 3.20<br>(0.22) | 2.28<br>(0.15) |
| Hours of wait before consultation. | 321.41 | 729.60<br>(49.46) | 6.95<br>(0.47) | 3.20<br>(0.22) | 6.84<br>(0.46) |
| Reduction in working hours (n=40) | 37.16 | 84.36<br>(5.72) | 2.10<br>(0.14) | 4.39<br>(0.30) | 0.00 |
| Days spent by relatives absent from work due to client illness (n=27) | 296 | 671.92<br>(45.55) | 24.88<br>(1.69) | 8.93<br>(0.61) | 18.15<br>(1.22) |
| Hours spent by relatives on transportation (n=46) | 134.59 | 305.52<br>(20.71) | 6.64<br>(0.45) | 3.91<br>(0.27) | 6.84<br>(0.46) |
| Direct Non-Medical Costs |  |  |  |  |  |
| Hours household spend on care for clients (n=36) | 214.94 | 487.92<br>(33.08) | 13.55<br>(0.92) | 8.80<br>(0.59) | 2.28<br>(0.15) |
| **Total** | **2,091.37** | **4,746.28**<br>**(321.78)** | **195.85**<br>**(13.28)** | **165.06**<br>**(11.19)** | **145.29**<br>**(9.85)** |

\*USD 1= GHS 14.75, interbank exchange rate, July 2024

The percentage distribution of costs associated with the treatment and management of BPH are as shown in figure 2 above. Direct medical costs constitute the largest proportion at 86.5%, largely surpassing direct non-medical costs, which account for 9.1%. These two categories together represent the vast majority (95.6%) of the total costs, highlighting the financial burden borne directly by individuals or households. In contrast, indirect costs, attributed to productivity losses, form a minimal share at 4.4%.

**Fig 2:**
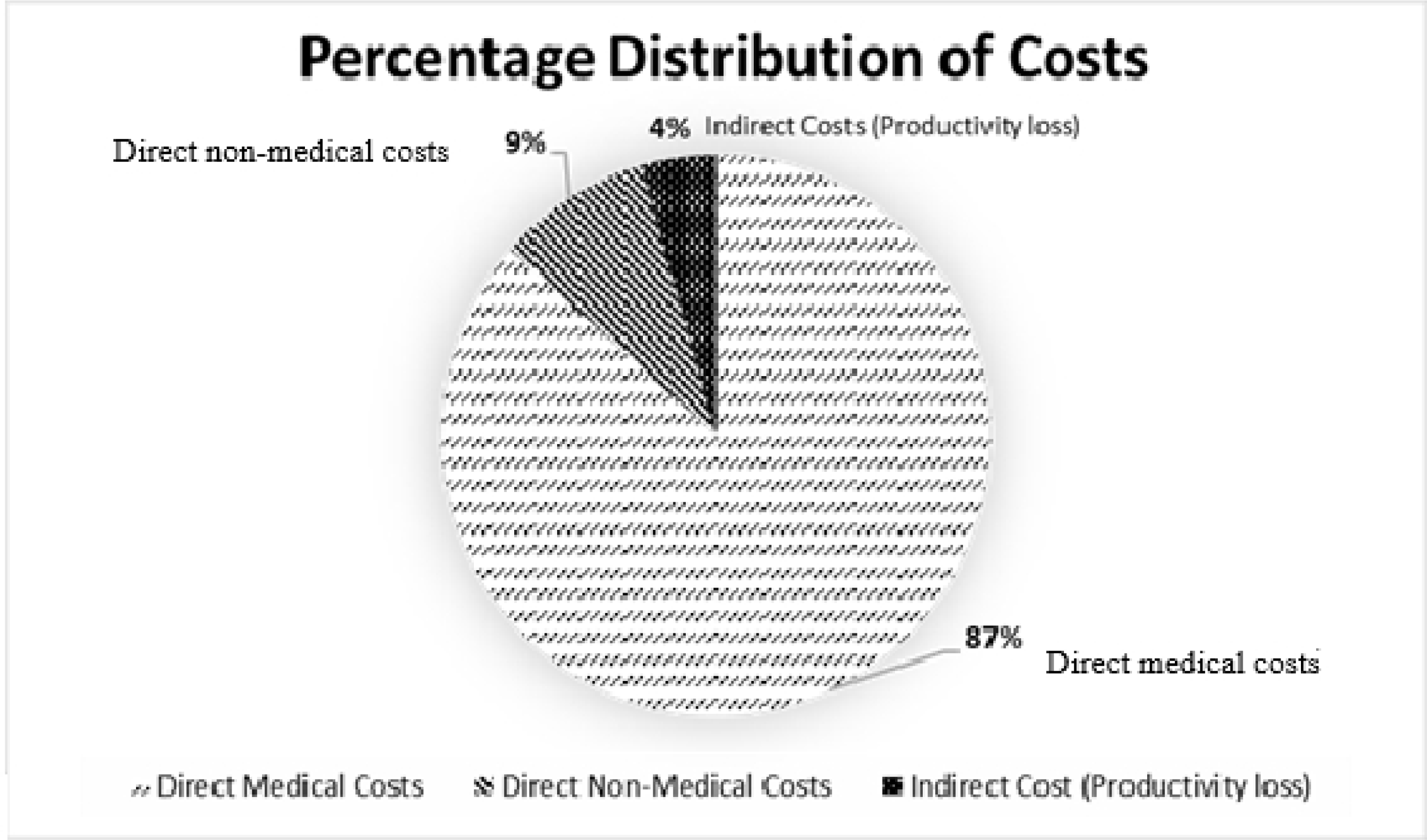
Percentage Composition of Costs.

## Discussion

The economic burden of diagnosing and treating BPH is considerable and varies based on symptom severity and healthcare setting. For instance, in Spain, annual costs for mild to severe symptoms ranged from €124 to €286, while in New Zealand, total direct medical cost for treatment was $NZ16 million. More recent studies by Ahn et al [10], highlighted the significant financial implications of long-term BPH management, especially with continuous medical therapy compared to early surgical intervention. In this study, it was found that the total direct monthly medical cost was GHS 93,462 ($5,964.87) with a mean cost of GHS 5,464.30 ($348.65) and a median cost of GHS 4,983.00 ($318.00). Diagnostic tests, including PSA, Transrectal ultrasound (TRUS), and uroflowmetry were associated with substantial costs, totaling GHS 27,044 ($1,725.54) with a mean cost of GHS 278.80 ($17.79) per patient. Medication costs amounted to GHS 21,078 ($1,345.67), with a mean of GHS 200.74 ($12.81), accounting for 22% of the total direct medical cost which is consistent with other studies indicating long term treatment with medication as a major financial burden [10]. Although medications are a major contributor to overall costs due to their long-term nature, surgeries have a higher per-patient cost. This is in line with the findings of Ahn et al [10], where early surgical intervention was noted to be a cost-effective option in the long term, despite the high upfront costs as typified by the relatively high cost of surgery in this study which accounted for 30% of the total direct medical cost output. Compared to findings from other studies conducted by Kovacs et al [8], costs of treatment in this study are lower, reflecting differences in healthcare pricing and availability of diagnostic services. Nonetheless, the financial burden on patients in Ghana is significant, especially given the lower average income levels and the out-of-pocket nature of healthcare expenditure. These findings underscore the need for cost-effective management strategies for BPH to alleviate the economic impact on patients.

The direct non-medical cost of BPH summed up to GHS 9,842.00 ($628.19), with an average cost of GHS 340.41 ± 175.83 ($21.72 ± $11.22) per patient. Transportation was the largest expense at GHS 6,523.00 ($416.24), averaging GHS 62.12 ± 96.62 ($3.97 ± $6.17) per patient, with variability attributed to wide differences in travel distance to and from the hospital facility, frequency of visits, and means of transport preferences. The median cost of transportation which is GHS 40.00 ($2.55) highlights that while many patients experience low transportation cost, others faced significant burdens, particularly those in rural areas far away from the hospital. Food and drinks were the second largest expense, amounting to GHS 3,029.00 ($193.19) with an average cost of GHS 33.29 ($2.12) per visit. Variations reflected differences in dietary needs, visit duration, and food availability. Lodging costs were negligible, with only one patient reporting an expense of GHS 200.00 ($12.76) as most patients were able to see a doctor on the same day of visit to the hospital. Usage of home care services was minimal as only two patients utilized the service at a total cost of GHS 90.00 ($5.74) per month. These findings align with studies, by Rencz et al [12] and Zou et al [13], where transportation was highlighted as a major cost for BPH patients. Although costs in the present study are lower, they are significant in the local economic context since the minimum wage in Ghana is significantly low (USD 1.16), emphasizing the need to address transportation and food expenses as critical components of the financial burden.

This study also highlights the significant economic and social burden of productivity losses associated with BPH treatment. Among 40 employed participants, 30% (12 participants) missed work and 27.5% reduced their working hours due to BPH. These figures are indicative of the pervasive impact BPH has on individuals’ ability to maintain steady employment. Global literature consistently underscores the disruptive nature of BPH, with studies by Garraway et al [16] revealing that BPH significantly interferes with daily activities, which often translates into lost productivity at work. In the context of Ghana, where formal employment is not always accessible and many individuals rely on daily wages or informal work, the impact of BPH can be even more pronounced. The total work hours lost amounted to 912 hours, translating to GHS 2,069.10 ($132.04) per month, with an average loss of GHS 137.94 ($8.80) per patient. Additionally, 15% of patients reported salary reductions, and 9.52% experienced job loss, further emphasizing the financial insecurity linked to BPH. The burden extended to caregivers, who experienced an estimated GHS 971.92 ($62.02) in productivity losses, with an average of GHS 24.88 ($1.59) per month, alongside additional caregiving and transportation costs. These findings are consistent with studies by Lambert et al [21], which noted the significant economic and emotional toll on caregivers. This study underscores the cascading effects of BPH on households, and overall quality of life in Ghana, where financial losses can have severe implications due to low average incomes as exemplified by mean hourly earnings of USD 1.01 in Ghana [22]. These findings emphasize the need for targeted interventions to reduce the economic and social burden of BPH, particularly in resource-constrained settings.

The cost distribution in Figure 2 reveals that direct medical costs constitute a substantial 86.5% of the total financial burden, consistent with findings from Rencz et al [12] and Ahn et al [10], which emphasize the significant impact of medication and physician visits on overall healthcare expenses. Notably, Ahn et al [10] reported that medication costs alone often exceed 50% of total treatment expenses in long-term medical therapy. In contrast to our study, which found direct non-medical costs to account for only 9.1% of the economic burden of BPH patients, Rencz et al [12] reported a higher proportion of 31%. This variation may be attributed to differences in transportation costs, accessibility of healthcare facilities, reliance on home care services, and lodging requirements. Additionally, indirect costs in our analysis were relatively low, contributing only 4.4%, compared to the 23% reported by Rencz et al [12], where productivity losses represented a more significant share of the financial burden. This discrepancy may stem from variations in healthcare systems, employment structures, and workplace policies governing individuals affected by BPH.

Our study relied on the minimum wage in the determination of the indirect cost output, this approach may have introduced bias since income variations occur across different sectors of the economy. Secondly, study estimates were based on data from a single institution thus the generalizability of findings may be limited. Thirdly, our study evaluated cost from the patients’ perspective; estimates from the societal or healthcare systems perspective are likely to be much higher. Nevertheless, these findings are worthwhile since they provide a baseline for future cost-of-illness studies on BPH in Ghana.

### Conclusion

The direct cost and productivity losses associated with BPH treatment had significant financial implications on patients and their households, with treatment modalities such as surgery and medications influencing the long-term cost of BPH. The average direct medical cost per patient was GHC 5,400 ($ 370), while transportation was the largest contributor to non-medical expenses. Productivity losses were substantial, as 30% of employed patients missed work, and 27.5% reported reduced working hours, leading to significant financial losses to patients and their households.

## Data Availability

The dataset underpinning this manuscript can be accessed at https://doi.org/10.6084/m9.figshare.28528601

## Acknowledgements

We are grateful to the participants of this study who provided useful information to evaluate the cost implications of prostate hyperplasia management in Ghana.

## Declarations

### Author contributions

DSDS conceptualized, collated data and wrote the draft manuscript, SGA, BK, FAA assisted in data collection and reviewed the draft manuscript, SKA reviewed the draft manuscript, YA was involved in data analysis and KKA reviewed the manuscript and supervised the project.

